# A pilot study: saliva oxytocin and testosterone in empathic stress responding

**DOI:** 10.1101/2025.11.28.25341194

**Authors:** Katja Heilmann, Martin Walter, Veronika Engert

**Affiliations:** Institute of Psychosocial Medicine, Psychotherapy and Psychooncology, Jena University Hospital, Friedrich Schiller University, Stoystr. 3, 07740 Jena, Germany; Clinic of Psychiatry and Psychotherapy, Jena University Hospital, Friedrich Schiller University, Philosophenweg 3, 07743 Jena, Germany; German Center for Mental Health (DZPG), parter site Halle-Jena-Magdeburg; Center for Intervention and Research in adaptive and maladaptive brain Circuits underlying mental health (C-I-R-C), Halle-Jena-Magdeburg, Germany

**Author notes:** Corresponding author: Katja Heilmann, Institute of Psychosocial Medicine, Psychotherapy and Psychooncology Jena University Hospital, Stoystr. 3, 07740 Jena.

**Keywords:** oxytocin, testosterone, empathic stress, stress contagion, vicarious stress, stress resonance

## Abstract

Suffering the stress of others (termed empathic stress or stress contagion) may affect an individual’s health and well-being. To understand the underlying hormonal processes of empathic stress, *N* = 108 opposite-sex dyads were tested, with one dyad partner passively observing the other undergo a standardized psychosocial laboratory stressor. A positive link between oxytocin release and empathic stress responding was hypothesized, while a negative relationship was expected for testosterone release. Associations of observer oxytocin and testosterone levels with two components of empathic stress were examined: stress resonance (i.e., synchronized observer - target responses) and vicarious stress (observer responses independent of target stress). During the passive observation of a stressed target, saliva oxytocin and testosterone levels increased by 16.64% and 23.00%, respectively, followed by a drop back to baseline levels. Supporting our hypotheses, increased observer oxytocin reactivity was linked to greater stress resonance in HF-HRV reactivity, while in low oxytocin responders, this association was reversed. Also, testosterone reactivity was negatively linked to stress resonance in heart rate activity. However, testosterone also showed a positive association with vicarious cortisol activity, which resembles the response pattern seen in first-hand stress exposure. We conclude that stress resonance may be the more empathy-dependent component of the empathic stress construct, aligning with previous work on the implication of oxytocin and testosterone in empathy. To draw further conclusions about the role of oxytocin and testosterone in empathic stress responding, future studies including a control group and stimulation of hormones are called for.

## 1 Introduction

Experiencing the stress of others is an increasingly documented phenomenon, referred to as “empathic stress” (also termed stress contagion, or stress attunement; Engert et al., 2019; Marheinecke et al., 2025; Nitschke and Bartz, 2023). Therein, without being exposed to the stressor itself, an observing individual experiences a “full-blown” physiological stress response by the mere observation of a stressed target individual. This is relevant, because repeated or prolonged exposure to empathic stress may negatively affect observers’ health and well-being (Engert et al., 2019), particularly when considering the impact of chronic stress exposure on the emergence of stress-related disease (Chrousos, 2009; Herman, 2022).

To date, the hormonal mechanisms underlying empathic stress occurrence remain unstudied (Engert et al., 2019; White and Buchanan, 2016). Therefore, we examined the link between observer oxytocin and testosterone release with their empathic stress response to an unknown target undergoing the Trier Social Stress Test (TSST; Kirschbaum et al., 1993), a standardized psychosocial laboratory stress test. Our study was conducted within a previous project investigating how observer’s empathic involvement (manipulated by prior first-hand stressor experience or being in a powerful position over the target) affected empathic stress (Heilmann et al., 2024).

Previous research in healthy humans suggests that empathic stress involves two components termed resonant and vicarious stress (Blasberg et al., 2023; Engert et al., 2019, 2014; Heilmann et al., 2024). In stress resonance, observers synchronize their response to that of the target. Observer and target stress activation are thus proportional to one another. Vicarious stress is independent of target stress and likely a projection of the observer perspective onto the target. In standardized laboratory stress paradigms, evidence for both components has been found in terms of hypothalamic-pituitary-adrenal (HPA) axis (specifically cortisol), parasympathetic and sympathetic nervous system activation (for reviews see Engert et al., 2019; Marheinecke et al., 2025; Nitschke and Bartz, 2023). Ecological validity of the laboratory-assessed results was demonstrated in one study showing that women who resonated more with their romantic partner’s cortisol release in the lab did likewise in everyday life (Engert et al., 2018b).

We and others proposed that empathy, the ability to infer and share another’s emotional state (de Waal and Preston, 2017), is essential for the emergence of empathic stress (Engert et al., 2019; Nitschke and Bartz, 2023). Indeed, observer’s trait and/or state empathy were mostly positively related to empathic stress (Blasberg et al., 2023; Buchanan et al., 2012; Engert et al., 2014; Pützer et al., 2020, but see Blons et al., 2021; Schury et al., 2020) or accelerated it (Dimitroff et al., 2017). Further factors modulating empathic stress include touch (Waters et al., 2017), emotional closeness (Blons et al., 2021; Engert et al., 2014; Manini et al., 2013), secure attachment (Gallistl et al., 2025), observation modality (live vs. video Engert et al., 2014), shared social identity (Schury et al., 2020, but see Erkens et al., 2019), social support, and proximity (Phan et al., 2019). Also, prior stressor experience reduced observers’ vicarious cortisol stress reactivity, while observers in a powerful position showed stronger vicarious autonomic stress responses (Heilmann et al., 2024). The role played by “social hormones”, such as oxytocin and testosterone, has not been investigated.

Oxytocin is a neuropeptide implicated in social behaviours such as affiliation, bonding (Insel and Young, 2001), social cognition and affect (Crespi, 2016; McCall and Singer, 2012). Accordingly, intranasal oxytocin administration in healthy humans was shown to increase compassion (Palgi et al., 2015; Rockliff et al., 2011), emotional (Hurlemann et al., 2010) and cognitive empathy (Bartz et al., 2019; Domes et al., 2007; Feeser et al., 2015; Stark et al., 2023). Similarly, self-reported empathy was positively associated with enhanced peripheral oxytocin levels after watching emotional videos (Barraza and Zak, 2009; Kampka et al., 2019; Procyshyn et al., 2020). Exceeding the role of a social hormone, oxytocin is additionally implicated in learning, memory, food intake, reproduction, aggression, and stress (Engert et al., 2016; Hoehne et al., 2022; Jurek and Neumann, 2018). Oxytocin is also sensitive to context, and released in context-specific anticipation (Bartz et al., 2011; Carter et al., 2020; Hoehne et al., 2022; Quintana and Guastella, 2020). Therefore, it was suggested to be an allostatic hormone with anticipatory features that regulates diverse behaviours to ensure the organism’s stability through changing environments (Quintana and Guastella, 2020). Considering oxytocin’s function in empathy, stress and its sensitivity to environmental cues, it is not surprising that in rodents, its involvement in stress contagion has already been confirmed (Peen et al., 2021). Here, intranasal oxytocin treatment enhanced freezing responses in mice observing distressed mice exposed to foot-shocks. Conversely, a systemic oxytocin receptor antagonist reduced freezing behaviour in the observing mice (Pisansky et al., 2017). Taken together, oxytocin is a promising marker to modulate the human empathic stress response.

Testosterone is a sex hormone (Eisenegger et al., 2011), mostly known for its role in social behaviours such as reproduction, aggression (Geniole and Carré, 2018), dominance (Mehta and Josephs, 2010), power (e.g., Stanton and Schultheiss, 2009) and social status (Eisenegger et al., 2011). It can display opposite effects to those of oxytocin, particularly in the context of affiliative behaviours (Crespi, 2016; McCall and Singer, 2012; Procyshyn et al., 2020, but see Reimers and Diekhof, 2015). For instance, testosterone administration in healthy humans can attenuate cognitive (Carré et al., 2015; Olsson et al., 2016; Van Honk et al., 2011, but see Nadler et al., 2019) and emotional empathy (Hermans et al., 2006). Further, higher endogenous testosterone levels were shown to relate to poorer empathic accuracy (Nitschke and Bartz, 2020; Ronay and Carney, 2013), as well as lower self-reported emotional and cognitive empathy (Chen et al., 2018, but see Zilioli et al., 2015). Notably, testosterone is responsive to psychosocial stress (Bedgood et al., 2014; Phan et al., 2017; Turan et al., 2015, but see Schoofs and Wolf, 2011), and its reactivity is highly depended on social contexts (Geniole and Carré, 2018; McCall and Singer, 2012). In sum, given its role in empathy, stress, and social contexts, testosterone is another potential player in empathic stress.

We therefore examined the relationship of oxytocin and testosterone release with empathic stress in *N* = 108 observers passively watching an unknown target perform the TSST through a one-way mirror. Based on the cited literature, we expected higher empathic (both resonant and vicarious) stress responses in subjective-psychological stress, cortisol, heart rate and high-frequency heart rate variability (HF-HRV) to be linked to increased oxytocin release. For testosterone, the opposite pattern was hypothesized. As each stress marker represents a different stress system of the multidimensional stress response (Engert et al., 2018a; Nater, 2018), hypotheses were analysed and interpreted separately for each marker (García-Pérez, 2023).

## 2 Materials and Methods

### 2.1 Participants

Between May 2021 and May 2022, 108 opposite-sex, stranger dyads from Jena and surroundings, Germany, enrolled as participants. Each dyad included a target undergoing the TSST and a passive observer. Of the 108 observers (age *M* ± *SD*: 24.74 ± 4.90, range: 19 – 39), 54 were female and observed a male target, while the other half was male and observed a female target (target sample: age *M* ± *SD*: 24.76 ± 4.79, range: 18 – 38). Only opposite-sex dyads were tested to prevent sex-specific influences on empathic stress responses and to provide comparability with previous empathic stress studies using opposite-sex dyads. Female participants were tested during their self-reported luteal phase to reduce variability due to menstrual cycle phase and to assure comparability with male cortisol responding (Kirschbaum et al., 1999) (for exclusion criteria and sample characteristics see the Supplemental Material).

The study was approved by the Research Ethics Boards of Friedrich-Schiller University Jena (ethic number: 2020-1643-BO), and performed in agreement with the Declaration of Helsinki. Participants provided written informed consent, were financially compensated, and could withdraw from the study at any time.

### 2.2 Design and testing procedure

Due to cortisol (Dallman et al., 2000) and testosterone circadian rhythms (Khan-Dawood et al., 1984), testing was conducted between 12 pm and 6 pm in one 150-min session at the Institute of Psychosocial Medicine, Psychotherapy and Psychooncology of Jena University Hospital. To standardize blood sugar levels, participants received a standardized snack upon arrival and were instructed to consume only water during testing. Participants were randomly assigned to their role (target vs. observer) and informed about it shortly after arrival, followed by 20 min of rest for laboratory adaptation. Electrocardiogram (ECG) breast belts were applied at -35 min prior to stressor onset (at 0 min). At -25 min, baseline subjective stress questionnaires and cortisol saliva samples were taken. Immediately thereafter, observers provided saliva samples for oxytocin and testosterone measurement.

After baseline sampling, observers were taken to the observation room, which was separated from the TSST room by a one-way mirror, and received instructions. They signed a two-line affirmation guaranteeing they would not need to perform the TSST themselves on that day to reduce potential stress induced by the situation. Subsequently, targets entered the TSST room, were instructed (at -15 min), and prepared for 10 min. At -3 min prior to stressor onset, stress questionnaires and cortisol samples were again collected. Through the one-way mirror, observers watched the target undergo the 10-min TSST stress phase (empathic TSST; Figure 1A). Afterwards, the third batch of subjective stress and saliva samples for all hormones were taken. Subsequently, targets and observers separately returned to their resting rooms. Throughout the 50-min recovery phase, subjective stress and hormone samples were collected, with the final samples taken at 60 min after stressor onset. Observers completed additional emotional state (at 15 min) and trait questionnaires (at 65 min) in randomized order (analysed and reported in Heilmann et al., 2024). The ECG device was removed at 50 min (see Figure 1B and 1C for the exact testing timeline). Targets were aware of being observed during their performance, but were always separated from observers.

**Figure 1.**
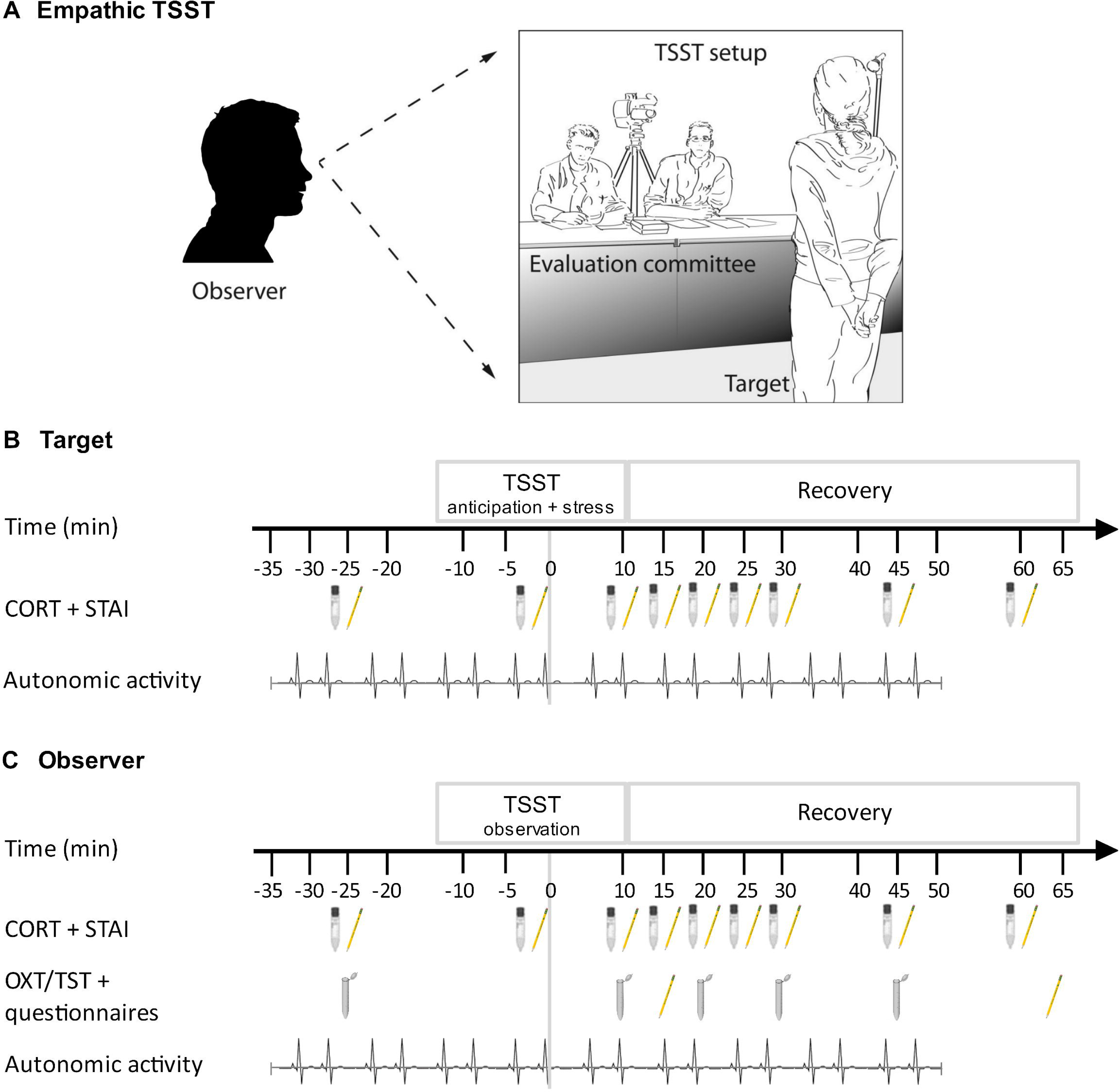
Study design: the empathic TSST (A) and the testing timeline for targets (B) and observers (C). (A) Targets were stressed with the Trier Social Stress Test (TSST, (Kirschbaum et al., 1993) while a opposite-sex observer watched through a one-way mirror. (B + C) Time is coded in minutes before and after stressor onset at 0 min. Salivary cortisol (CORT) was assessed via Salivettes, and subjective stress via the state scale of the State Trait Anxiety Inventory (STAI, Spielberger et al., 1983). In observers, salivary oxytocin (OXT) and testosterone (TST) were collected with Salicaps after the collection of cortisol. An electrocardiogram for the assessment of autonomic activity (heart rate and high-frequency heart-rate variability) was recorded from -35 to 50 min relative to stressor onset. Observers completed emotional state response scales (at 15 min) and trait questionnaires (at 65 min after stressor onset; analysed and reported in Heilmann et al., 2024).

Since the pilot study was conducted within another project, 72 observers underwent a manipulation which either required them to perform the TSST as targets themselves prior to the empathic TSST (*n* = 36) or to take a position of power by deciding on an alleged additional financial compensation for the target (*n* = 36; see Heilmann et al. (2024) for a detailed description and effects of the manipulation).

### 2.3 Stress induction

The Trier Social Stress Test (TSST; Kirschbaum et al., 1993) was used to induce psychosocial stress in targets, while observers watched through a one-way mirror (empathic TSST; Figure 1A). It involved a 15-min anticipation phase, a 5-min audio- and videotaped mock job talk, and a 5-min difficult mental arithmetic task. Both tasks were completed before a sex-mixed, non-empathic committee of two alleged behavioural analysts.

### 2.4 Measures

#### 2.4.1 Salivary cortisol

Saliva cortisol as marker of HPA axis activity (Miller et al., 2013) was simultaneously collected at -25 min, -5 min, 10 min, 15 min, 20 min, 25 min, 30 min, 45 min, and 60 min relative to stressor onset in observers and targets using Salivettes (Sarstedt, Nümbrecht, Germany; Figure 1B and 1C). Participants placed the Salivette in their mouth, and refrained from chewing for 2 min. Salivettes were stored at -80°C until assay at the Biochemical Laboratory of the Department of Biological and Clinical Psychology, Trier University. For determination of cortisol levels (nmol/l), a time-resolved fluorescence immunoassay with intra- and interassay variabilities of less than 10% and 12% was used (Dressendörfer et al., 1992). Samples were assayed in duplicate.

#### 2.4.2 Salivary oxytocin and testosterone

Observers’ salivary oxytocin and testosterone were assessed at -25 min, 10 min, 20 min, 30 min, and 45 min relative to stressor onset by passive drool into 2 ml Salicap collection devices (IBL International GmbH; Figure 1C). Samples were stored at -80°C until assay at the Institute of Medical Psychology, Heidelberg University Hospital. Oxytocin levels (pg/ml) were determined using a competitive enzyme immunoassay, the Oxytocin ELISA kit (ADI-901-153A, Enzo Life Sciences), with intra- and interassay variabilities of 6.31% and 10.34%. Oxytocin samples were not extracted, as saliva oxytocin concentrations seem to be highly correlated regardless of whether or not sample extraction is performed (MacLean et al., 2018). For determination of testosterone levels (pg/ml), a luminescence immunoassay (RE62031 / RE62039, IBL International GmbH) with intra- and interassay variabilities of 3.8% and 8.25% was used. Fifty percent of samples were assayed in duplicate.

#### 2.4.3 Heart rate and high-frequency heart rate variability

Heart rate as a marker of sympathetic (Grassi et al., 1998) and high-frequency heart rate variability (0.15 -0.4 Hz; HF-HRV) as a marker of parasympathetic nervous system (vagus nerve) activity were assessed (Berntson et al., 1997; The Task Force of the European Society and The North American Society of Pacing and Electrophysiology, 1996). Heart rate and HF-HRV were simultaneously recorded for 85 min (-35 to 50 min relative to stressor onset; Figure 1B and 1C) in observers and targets by a continuous ECG using the Zephyr Bio-Harness 3 (Zephyr Technology, Annapolis, Maryland, USA) and a sample frequency of 250 Hz (for timing and duration of the ECG time frames see Table S2 in the Supplemental Material).

Using an in-house created Python-based script (Python Language Reference, 2018), raw recordings were automatically corrected and manually checked for artifacts by two independent researchers. For each time frame, average HR (beats per minute; bpm) and HF-HRV (square milliseconds; ms^2^) according to the Lomb-Scargle periodogram were calculated using the “hrv-analysis 1.0.4” Python package (Champseix et al., 2021).

#### 2.4.4 Subjective stress experience

Subjective stress experience was simultaneously assessed in targets and observers (at - 25 min, -5 min, 10 min, 15 min, 20 min, 25 min, 30 min, 45 min, and 60 min relative to stressor onset; Figure 1B and 1C) using the 20-item state scale of the State Trait Anxiety Inventory (STAI) (Spielberger et al., 1983). The STAI asks for feelings of apprehension, nervousness, tension, worry, and activation/arousal of the autonomic nervous system (Spielberger et al., 1983) (for further state and trait measures see the Supplemental Material).

### 2.5 Statistical analyses

Initially, the current analyses were planned as a joint project with those published in (Heilmann et al., 2024), and preregistered together (https://osf.io/nbspw). However, to not overload the resulting manuscript, we decided to publish the oxytocin and testosterone data separately. While this naturally leads to some deviation, we stayed as close to the preregistration as possible (see the Supplementary Materials for details). The analysis script is accessible at https://osf.io/uxed2/. The dataset generated and/or analyzed for the current study are not publicly available due to ongoing analyses in the project. Data are available upon reasonable request.

As this pilot study was part of a greater project, power analysis could not be conducted for the individual study. However, our sample size (*N* = 108 dyads) matches or exceeds samples sizes of previous studies analysing oxytocin and/or testosterone data in empathy research (Barchi-Ferreira and Osório, 2021; Carré et al., 2015; Nitschke and Bartz, 2020; Stark et al., 2023).

Analyses were performed with the software R, version 4.2.2 (R Core Team, 2020). Physiological markers (cortisol, heart rate, HF-HRV, oxytocin and testosterone) were ln-transformed to account for skewness (see the Supplemental Material for missing replacements and dealing with outliers). Significance level was set to *p* ≤ 0.05. The grouping variable of the manipulation and observer sex were set as fixed covariates. Additional covariates (observer age, BMI, menstrual cycle status, education level) were included only when increasing model fit.

#### 2.5.1 Descriptive Statistics

Targets and observers exhibiting a minimal cortisol increase of 1.5 nmol/l from baseline to their individual peak were classified as cortisol responders (Miller et al., 2013). Observers’ average increases in oxytocin and testosterone levels from baseline to their individual peak were calculated and reported in percentage.

To verify statistical significance of observer oxytocin and testosterone release, two LMMs with either oxytocin or testosterone data as dependent variables were calculated. Repeated measures of oxytocin and testosterone were nested within individuals (observers). Of the five oxytocin measurements, only the first three were used because, after rising and dropping back to baseline levels, oxytocin started to rise again (at 32 min after stressor onset), indicating a confounding influence outside the empathic TSST (Figure 2A). To model the time course of each hormone (i.e., observer reactivity and recovery), two continuous time variables were created. The intercept was defined as the measurement time point of observers’ highest average peak (at 12 min after stressor onset for both hormones) and set to 0. The two continuous time variables modelled the minutes between measurement time-points: from baseline to peak (reactivity slope: -23 min to 12 min relative to stressor onset) and from peak to recovery (recovery slope: 12 to 22 min for oxytocin and 12 to 47 min for testosterone). Observer sex, the manipulation group variable, and observer age (only for the testosterone model) were included as covariates. Contrasts were set to the predefined “contr.sum” in R, so that our intercept represented the grand mean of the sample and not the mean of one category in the categorical covariates.

**Figure 2.**
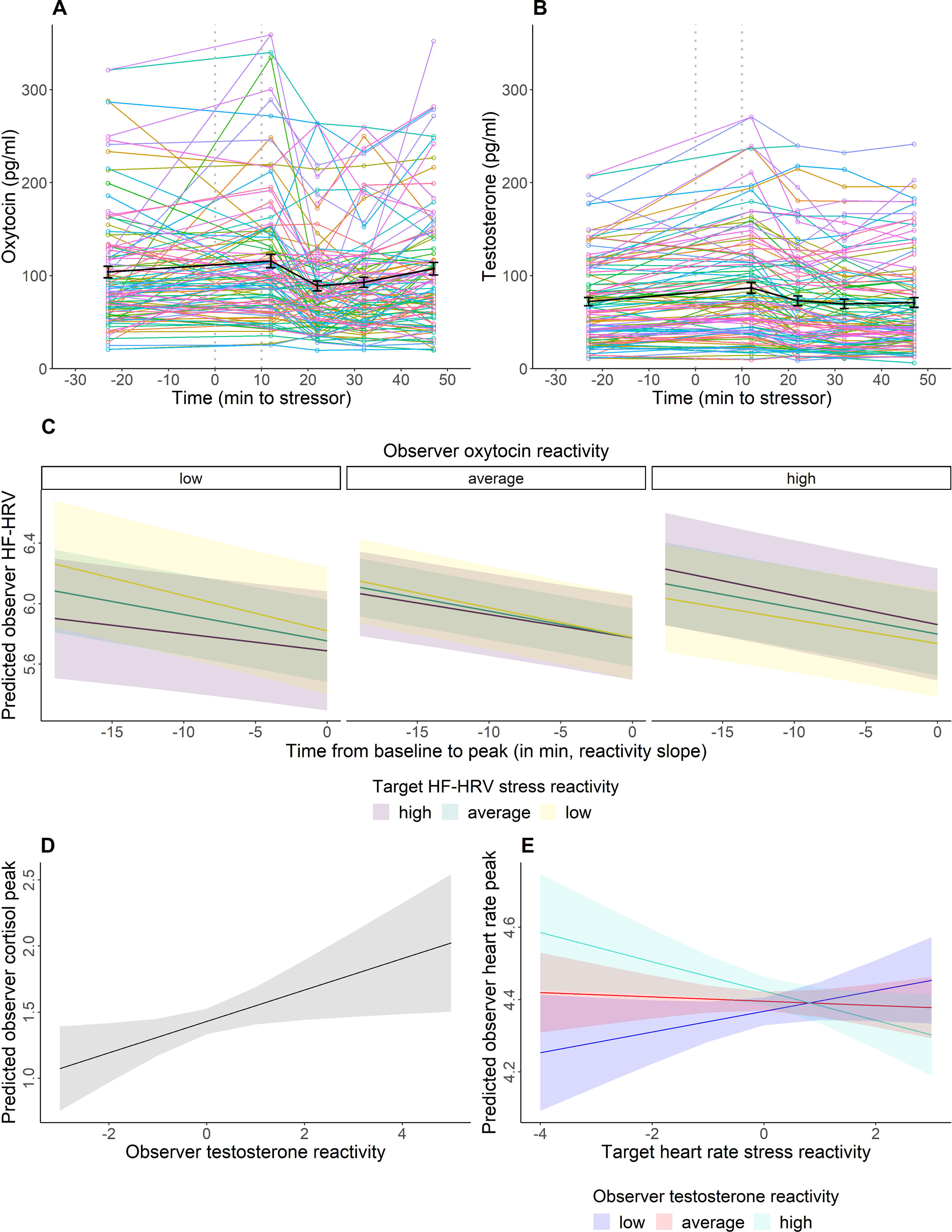
**A, B: Individual trajectories of observer oxytocin (A, raw data) and testosterone levels (B, raw data) during the empathic stress paradigm**. In black, mean oxytocin and testosterone levels including standard errors as error bars are given. Time is given relative to stressor onset. Data were winsorized to 3 *SDs* to the mean. Because covariates are not considered, results may deviate from the model-derived depiction. **C, D, E: Model-derived predictions of associations between observer oxytocin reactivity with stress resonance in high-frequency heart rate variability (HF-HRV) reactivity, (C) and of associations between observer testosterone reactivity with the cortisol peak (D) and stress resonance in the heart rate peak (E).** (C) Observers with a relatively low oxytocin reactivity to the empathic TSST revealed stronger HF-HRV stress reactivity (i.e., a steeper reactivity slope), when targets were low stressed and vice versa (C, left). In observers exhibiting an average oxytocin response to the empathic TSST, the targets’ stress reaction had no effect on their HF-HRV stress reactivity (C, middle). Observers having relatively high oxytocin reactivity revealed HF-HRV stress resonance. That is, a steeper HF-HRV reactivity slope when observing a highly stressed target (C, right). (D) The more the testosterone reactivity of observers increased during the empathic TSST, the higher was their vicarious cortisol peak (indicating a vicarious cortisol response to the entire testing situation rather than just the TSST). (E) The more the observer testosterone reactivity to the empathic TSST increased, the less pronounced was their stress resonance in heart rate peak (indicating a resonant response to the entire testing situation rather than just the TSST), up to a reversal of the effect in highly testosterone reactive observers. 95% confidence intervals are given. All data were winsorized to 3 *SDs* to the mean. Cortisol (nml/l), heart rate (bpm), and HF-HRV (ms^2^) data were logerithmized. Target stress and observer testosterone/oxytocin reactivity were z-standardized. For Figure C, time is depicted relative to the average peak time frame (from 5 to 10 min after stressor onset), which was set to 0 in the models.

#### 2.5.2 The role of oxytocin and testosterone in empathic stress responses

Associations of observer oxytocin and testosterone reactivity with empathic stress responses in subjective stress, cortisol, heart rate and HF-HRV were tested with LMMs (adding up to four models for oxytocin and testosterone each). Repeated measures of a stress marker were nested within individuals (observers). The intercept, defined as the measurement time point of observers’ highest average stress-induced peak, was set to 0. To estimate the empathic stress reactivity of observers for a given marker, a continuous time variable modelled the minutes between measurement time-points from baseline to peak (reactivity slope; see Table S4 for the exact timeline of each marker). Contrasts were set to the predefined “contr.sum” in R, so that our intercept represented the grand mean of the sample. Due to this statistical approach, the peak (model intercept) was independent of stress reactivity, and interpreted as the level of overall activation per stress marker. Target stress reactivity scores (Δtarget), observer oxytocin or testosterone reactivity scores (Δoxytocin or Δtestosterone), their interactions with the reactivity slopes, and covariates (observer sex and the manipulation group variable for all models; observer age only for the HF-HRV model) were added to the model. ΔTarget, observer Δoxytocin and Δtestosterone were operationalized as a change score from baseline to the individual peak. Physiological change scores were further adjusted for baseline levels by extracting the standardized change score residuals from a regression model. Since inferences were drawn separately for each stress marker and hormone, p-values were not corrected for multiple testing (García-Pérez, 2023).

Significant main effects of the intercept (peak) and reactivity slope represented vicarious stress. Resonant stress responses, being dependent on the target reactivity, were represented by a main effect of target stress reactivity (Δtarget), and an interaction of Δtarget with the reactivity slope. Associations of observer oxytocin or testosterone reactivity with vicarious stress (overall levels: main effect of Δoxytocin/Δtestosterone; reactivity: reactivity slope*Δoxytocin/Δtestosterone interaction) and resonant stress (overall levels: Δtarget*Δoxytocin/Δtestosterone; reactivity: reactivity slope*Δtarget*Δoxytocin/Δtestosterone) were tested (see the Supplemental Material for model equations and further statistical details).

## 3 Results

### 3.1 Descriptive statistics

A physiologically significant cortisol increase of 1.5 nmol/l from baseline (Miller et al., 2013) was found in 75% of the targets, indicating successful stress induction (Kudielka et al., 2007). Increases in cortisol of ≥ 1.5 nmol/l were also seen in 14.95% of observers, aligning with empathic stress studies in strangers (10 to 17% in Engert et al., 2014; Erkens et al., 2019; Schury et al., 2020). Oxytocin levels rose on average by 16.65% and testosterone levels by 23.00% from baseline to peak. In support of this, LMMs examining observer oxytocin and testosterone release revealed that levels of both hormones increased during the empathic TSST (main effect of reactivity slope, oxytocin: β = 0.003, 95% CI [0.001; 0.005], *t*_208_= 2.41, *p* = .017, testosterone: β = 0.002, 95% CI [0.001; 0.004], *t_430_*= 2.99, *p* = .003), and dropped back to baseline levels (main effect of recovery slope, oxytocin: β = -0.025, 95% CI [-0.033; - 0.017], *t*_209_= -5.96, *p* < .001, testosterone: β = -0.006, 95% CI [-0.007; -0.004], *t_430_*= -6.69, *p* < .001; Table 1).

**Table 1.** A LMM examining observer oxytocin (A) and testosterone levels (B) during the empathic stress paradigm.

| Fixed effects | A Oxytocin |  |  | B Testosterone |  |  |
| --- | --- | --- | --- | --- | --- | --- |
| | $\beta$ | 95% CI | $t$ (df) | $\beta$ | 95% CI | $t$ (df) |
| Intercept (peak) | 4.590 | 4.484; 4.697 | 83.35 (151)*** | 4.133 | 4.049; 4.216 | 95.37 (129)*** |
| Reactivity slope | 0.003 | 0.001; 0.005 | 2.41 (208)* | 0.002 | 0.001; 0.004 | 2.99 (430)** |
| Recovery slope | -0.025 | -0.033; -0.017 | -5.96 (209)*** | -0.006 | -0.007; -0.004 | -6.69 (430)*** |
| Sex | 0.090 | -0.006; 0.187 | 1.82 (102)† | 0.603 | 0.524; 0.683 | 14.62 (103)*** |
| Group1 | -0.097 | -0.233; 0.040 | -1.37 (102) | -0.042 | -0.154; 0.069 | -0.74 (103) |
| Group2 | -0.097 | -0.233; 0.038 | -1.39 (102) | -0.138 | -0.250; -0.027 | -2.39 (103)* |
| Age |  |  |  | -0.102 | -0.179; -0.021 | -2.45 (103)* |
| **Random Effects** |  |  |  |  |  |  |
|  | Variance (*SD*) |  |  | Variance (*SD*) |  |  |
| Individual | 0.23 (0.48) |  |  | 0.17 (0.41) |  |  |

*Note.* Satterthwaite approximation for degrees of freedom was used. Models based on 318 observations from 107 dyads (A, oxytocin) and 540 observations from 108 dyads (B, testosterone). $\beta$ = standardized beta coefficients; CI = confidence interval. Contrasts were set to the predefined “contr.sum” in R, so that the intercept (peak) represented the grand mean. Oxytocin and testosterone data were winsorized to 3 *SDs* to the mean and logerithmized. Age was z-standardized.
† $p < .10$ , \* $p < .05$ , \*\* $p < .01$ , \*\*\* $p < .001$ .

### 3.2 The role of oxytocin and testosterone in empathic stress

Independent of the oxytocin and testosterone results, the LMMs confirmed the empathic stress results reported in Heilmann et al. (2024). In short, vicarious stress reactivity emerged in subjective stress and HF-HRV, but not in cortisol and heart rate. Further, target stress reactivity predicted observers’ overall cortisol release throughout the testing session, indicating higher cortisol release across the session when observing highly stressed targets (all statistical parameters are summarized in Table 2 and 3).

**Table 2.**
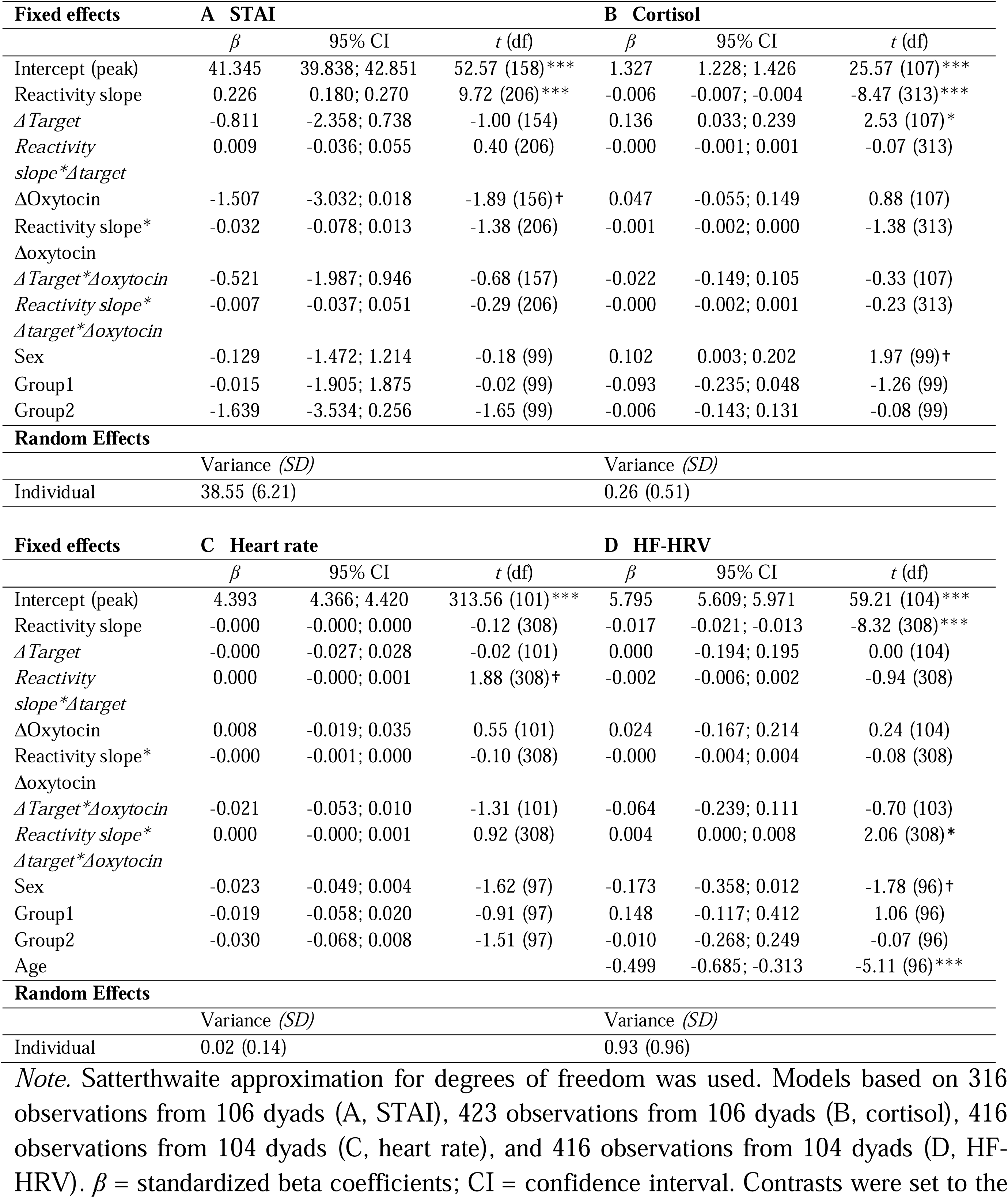
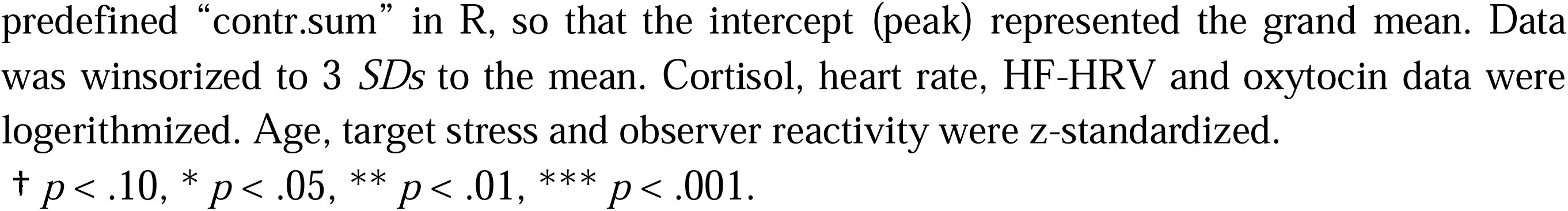
A LMM examining associations between observer oxytocin reactivity and empathic stress responses in the state scale of the State Trait Anxiety Inventory (STAI, Spielberger et al., 1983; A), cortisol (B), heart rate (C) and high-frequency heart rate variability (HF-HRV; D). Predictor variables written in italics represent resonant stress responses (dependent on target’s stress). ΔTarget: target stress reactivity; Δoxytocin: observer oxytocin reactivity.

**Table 3.**
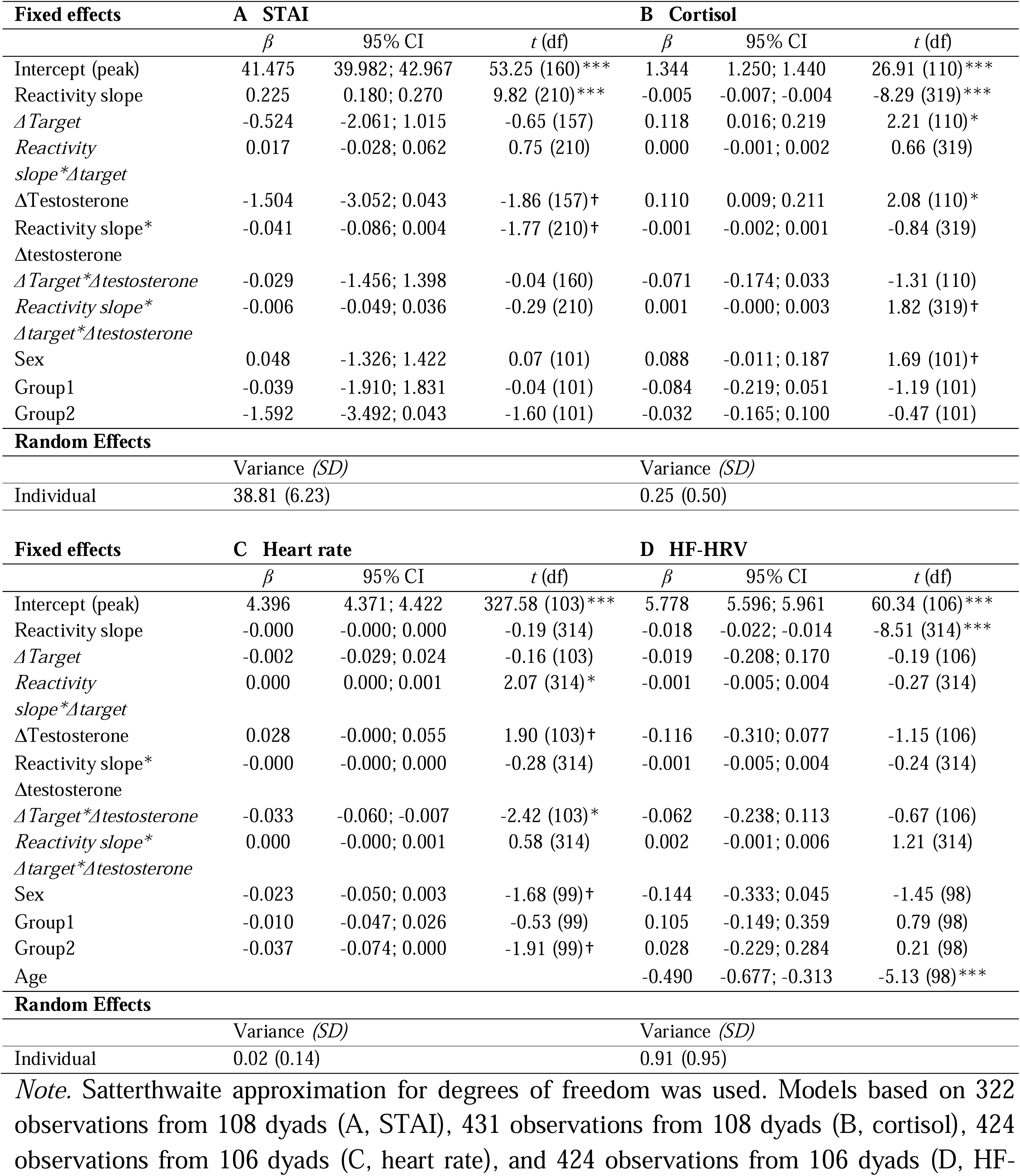
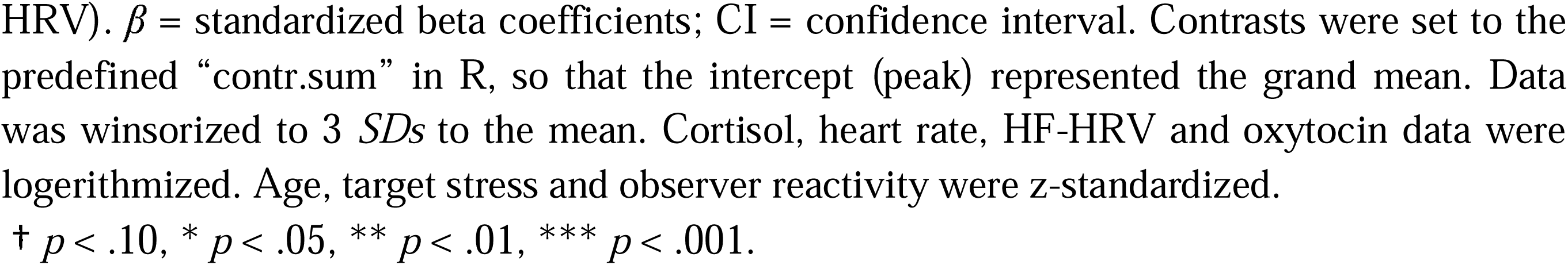
A LMM examining associations between observer testosterone reactivity and empathic stress responses in the state scale of the State Trait Anxiety Inventory (STAI, Spielberger et al., 1983; A), cortisol (B), heart rate (C) and high-frequency heart rate variability (HF-HRV; D). Predictor variables written in italics represent resonant stress responses (dependent on target’s stress). ΔTarget: target stress reactivity; Δtestosterone: observer testosterone reactivity.

#### 3.2.1 Associations of observer oxytocin reactivity with empathic stress responses

No associations of observer oxytocin reactivity with vicarious stress reactivity (i.e., observer stress responses irrespective of target stress) were found across stress markers (reactivity slope*Δoxytocin: all *t* ≤ |1.38|, all *p* ≥ .170, Table 2).

Concerning stress resonance (i.e., synchronization of observer and target stress responses), there was a link between oxytocin reactivity and HF-HRV reactivity (reactivity slope*Δtarget*Δoxytocin: β = 0.004, 95% CI [0.000; 0.008], *t*_308_= 2.06, *p* = .041; Table 2D), such that higher oxytocin release was related to higher HF-HRV stress resonance (i.e., a steeper reactivity slope when observing a highly stressed target; Figure 2C, right). In contrast, observers exhibiting lower oxytocin reactivity showed the opposite pattern with greater HF-HRV stress reactivity when confronted with a low-stress target (Figure 2C, left). No significant effects were found for subjective stress, cortisol, and heart rate (all *t* ≤ |0.92|, all *p* ≥ .358, Table 2).

#### 3.2.2 Associations of observer testosterone reactivity with empathic stress responses

Again, no associations emerged between testosterone *reactivity* and vicarious stress reactivity across markers (reactivity slope*Δtestosterone: all *t* ≤ |1.77|, all *p* ≥ .079, Table 3). However, testosterone reactivity was associated with *overall* cortisol levels (operationalized as the peak and indicating a vicarious response to the entire testing situation rather than just the TSST; main effect of Δtestosterone: β = 0.110, 95% CI [0.009; 0.211], *t*_110_= 2.08, *p* = .040; Table 3B). Specifically, observers with higher testosterone reactivity during the empathic TSST exhibited higher overall cortisol release throughout the testing session (Figure 2D).

Stress resonance *reactivity* was unrelated to testosterone reactivity across stress markers (reactivity slope*Δtarget*Δtestosterone: all *t* ≤ |1.82|, all *p* ≥ .070, Table 3). However, there was a link between testosterone reactivity and *overall* heart rate activity (operationalized as the peak and indicating a resonant response to the entire testing situation rather than just the TSST; Δtarget*Δtestosterone: β = -0.033, 95% CI [-0.060; -0.007], *t*_103_= -2.42, *p* = .017; Table 3C). In detail, given reduced testosterone reactivity during the empathic TSST, observers’ heart rate activity was higher throughout the testing session when witnessing highly stressed targets. Conversely, given increased testosterone reactivity in observers, this effect reversed (Figure 2E).

## 4 Discussion

In aiming to understand the hormonal mechanisms underlying empathic stress occurrence, we explored associations of oxytocin and testosterone release with empathic stress responses in observers while they passively watched an unknown, opposite-sex target performing a standardized psychosocial laboratory stressor (Trier Social Stress Test, TSST; Kirschbaum et al., 1993). Two components of empathic stress were investigated for subjective stress, cortisol, heart rate, and HF-HRV: vicarious stress (observer reactivity independent of target stress reactivity) and resonant stress (observer reactivity proportional to target reactivity).

Despite not being exposed to the TSST themselves, 14.95% of observers showed physiologically significant cortisol release of at least 1.5 nml/l when witnessing a target undergo first-hand stress. Further, an average rise of 16.64% in saliva oxytocin and of 23.00% in saliva testosterone levels was detectable in observers, followed by a drop back to baseline. Increased observer oxytocin reactivity was related to stronger resonance in HF-HRV reactivity, while reduced oxytocin reactivity was associated with the reversed effect (i.e., stronger HF-HRV reactivity when observing less stressed targets, Figure 2C). Testosterone reactivity was positively linked to vicarious cortisol release throughout the testing, but negatively to resonance in overall heart rate activity (Figure 2D and E).

Intriguingly, peripheral levels of oxytocin and testosterone in observers increased and dropped back to baseline levels while they were passively observing a stressed target (Figure 2). As no control group was included, it is unclear whether these findings can be exclusively attributed to the observation of a stressed target or are caused by the experimental setting itself (e.g., coming to the lab or interacting with the research assistant). However, three arguments would suggest specificity to the empathic TSST. First, both, saliva oxytocin and testosterone release, are sensitive (i.e., increase in response) to first-hand TSST exposure (Bedgood et al., 2014; de Jong et al., 2015; Engert et al., 2016; Hoehne et al., 2022; Phan et al., 2017). Hence, it is reasonable to assume that also second-hand exposure to the TSST might lead to rises in saliva oxytocin and testosterone levels. Second, the empathic TSST accounted for most of the time between baseline and peak sampling (15 min TSST anticipation and 10 min TSST performance). In other words, there was not much else experimental content to react to. Third, observing a stressed target was possibly a much more emotionally salient stimuli than other activities within this time frame which were not directly linked to the experience of empathic stress (such as signing a two-line affirmation or listening to instructions). Therefore, it is most likely that the average rise and fall of oxytocin and testosterone levels was triggered by observing the stressed target. Given this assumption, our findings could suggest that peripheral oxytocin and testosterone might be sensitive to an empathic stress context, potentially due to the situation’s social and evolutionary relevance. Learning from others’ stressful experiences may be adaptive and preparatory if confronted with similar, future stressors, suggesting an adaptive role for both hormones (de Vignemont and Singer, 2006; McCall and Singer, 2012; Quintana and Guastella, 2020).

Our main focus was on whether oxytocin and testosterone were involved in empathic stress responding. Specific assumptions were that higher observer oxytocin and lower testosterone reactivity would relate to higher vicarious and resonant stress responses in subjective stress, cortisol, heart rate, and HF-HRV.

While observer oxytocin reactivity was not associated with vicarious stress responses during the empathic TSST, we found higher resonance in HF-HRV reactivity with increasing oxytocin release (Figure 2C). Hence, supporting our hypothesis, higher oxytocin release was positively linked to parasympathetic stress resonance. With decreasing oxytocin release, this association reversed, such that parasympathetic reactivity in observers was lower with higher target stress. One can speculate that with lower oxytocin release, observers might have perceived the empathic stress situation as less salient, leading to lower (or even inversed) resonance with the stressed targets. With an increasing number of empathic stress studies available, it is becoming apparent that parasympathetic activation seems to be particularly prone to resonance effects, possibly due to its role in threat appraisal (Thayer et al., 2012) and social processes (Porges, 2021). Thus, the fact that only parasympathetic resonance was linked to oxytocin reactivity adds to findings in mother-child dyads where particularly HF-HRV resonance was boosted by emotional mimicry in adolescents (preprint: Blasberg et al., 2024), and by theory of mind capacity in children (Blasberg et al., 2023).

Concerning testosterone, higher reactivity during the empathic TSST was linked to increased vicarious cortisol release throughout the testing session. In other words, more stress-sensitive observers, exhibiting higher cortisol levels already prior to the empathic TSST, additionally released more testosterone during the empathic stress situation. Although contrary to our hypothesis, these results align with findings showing a coupling of testosterone and cortisol release in response to first-hand experienced social evaluative stressors like the TSST (Knight et al., 2017; Phan et al., 2017; Shirtcliff et al., 2015; Turan et al., 2015). Importantly, Phan et al. (2017) not only found a coupling in *reactivity*, but also in *overall* levels of testosterone and cortisol throughout the testing. As suggested by Turan et al. (2015), this coordinated response may be adaptive in status-challenging situations, with testosterone enhancing performance and cortisol supplying energy resources. Toward that extent, the TSST may be perceived as a stressor and challenge (Phan et al., 2017), not just when experienced first-hand, but also when passively observed.

For stress resonance, a pattern opposite to that of vicarious stress was found. Specifically, a negative association of testosterone reactivity with overall heart rate resonance indicated that, given decreased testosterone reactivity, heart rate activity throughout the testing session was increased if targets were highly stressed. Like the association between oxytocin release and HF-HRV resonance, this effect reversed for highly testosterone-reactive observers. Thus, supporting our hypothesis and as suggested by other empathy studies (e.g., Carré et al., 2015; Chen et al., 2018; Hermans et al., 2006; Nitschke and Bartz, 2020; Olsson et al., 2016), improved ability to tune in to the state of a stressed target was related to reduced testosterone reactivity during the empathic TSST. Like the oxytocin findings, this effect was limited to a marker of autonomic activity, although not HF-HRV. Other than expected, the current association was found in overall levels of heart rate activity throughout the testing session rather than in reaction to the empathic TSST. An explanation may be that observers who were sympathetically aroused by the testing situation right from the start relocated their attention to particularly salient cues (i.e., the stressed targets; Hermans et al., 2014). Consequently, their empathic accuracy to assess the targets’ stressful experience may have been improved (Nitschke et al., 2022), leading to resonance in overall activity rather than in reactivity.

Oxytocin and testosterone showed distinct association patterns with vicarious and resonant stress, and only for resonant stress, results aligned with previous empathy studies (i.e., showing more empathy with higher oxytocin and lower testosterone release). The association of testosterone with higher vicarious cortisol activity rather mirrored a typical firsthand stress response. Therefore, although resonant and vicarious stress relate to self-reported empathy (Blasberg et al., 2023; Buchanan et al., 2012; Engert et al., 2014), we suggest that stress resonance may require greater empathic ability than vicarious stress occurrence. Considering the role of oxytocin in empathy (Barchi-Ferreira and Osório, 2021; Hurlemann et al., 2010; Stark et al., 2023), this might explain why an association with oxytocin reactivity was found only in stress resonance.

Several study limitations need to be considered. First, a control group with no exposure to the empathic TSST was missing, limiting the interpretability of our findings. Second, associations were investigated, forbidding to draw causal conclusions about the involvement of oxytocin and testosterone in empathic stress. Moreover, as only peripheral (saliva) levels of oxytocin and testosterone were measured, the results do not apply to their (central) levels in the brain, although, for oxytocin, there has been evidence suggesting that peripheral and central levels in the brain align under periods of stress (Valstad et al., 2017). Further, despite statistically controlling for its influence, the observer manipulation of either prior stressor experience or being in a powerful position may have influenced results. Regarding oxytocin and testosterone saliva sampling, some participants indicated to perceive sampling as unpleasant and slightly stressful. Also, participants needed varying amounts of time (2 to 10 min) to provide sufficient saliva. Finally, generalizability might have been reduced due to the young, healthy, and culturally homogenous sample.

In summary, saliva oxytocin and testosterone levels increased and dropped back to baseline levels while observing an unknown target undergoing psychosocial stress. Despite their similar responsivity, their associations with empathic stress responses showed distinct and even opposite patterns. Increased observer oxytocin reactivity was related to higher stress resonance in HF-HRV. Conversely, higher testosterone reactivity was linked to reduced stress resonance in overall heart rate, and to increased overall vicarious cortisol activity. Testosterone’s role in vicarious stress mirrored its role in first-hand stress. Associations in stress resonance, however, were in accordance with empathy studies, suggesting stress resonance as the more empathy-dependent component of empathic stress. We encourage future studies using a control group and exogeneous stimulation to uncover the casual role of oxytocin and testosterone in empathic stress.

## Supporting information

Supplements_Heilmann

## Data Availability

All data produced in the present study are available upon reasonable request to the authors

## 6 Declarations

## Acknowledgements

We are thankful to the members of the Social Neuroscience Department and Social Stress and Family Health Research Group involved in the *HOPES* project in particular to Henrik Grunert for technical assistance and to all research assistants and students, whose help with data collection was indispensable.

## 7 Author Statement

Author contributions: K.H. and V.E. developed and designed the experiment. K.H. conducted data collection, preparation, and analyses under the supervision of V.E.. K.H. drafted, and all authors critically revised and approved the manuscript.

