## Supplements_Heilmann for "A pilot study: saliva oxytocin and testosterone in empathic stress responding"

### saliva oxytocin and testosterone in

### empathic stress responding

[*]Corresponding author:

Katja Heilmann

Jena University Hospital

Stoystr. 3, 07740 Jena

### Supplementary Material

#### This PDF file includes

#### Supplementary Materials and Methods

##### Supplementary exclusion criteria and sample characteristics

Exclusion criteria covered pregnancy, menopause, an abnormal body mass index (BMI; BMI < 18 or BMI > 30), regular smoking (> 10 cigarettes a day) and drug use, all due to their influence on HPA axis regulation (Incollingo Rodriguez et al., 2015; Kajantie and Phillips, 2006; Packard et al., 2016; Rohleder and Kirschbaum, 2006). Further, participants self-reporting recent stressful life events (e.g., loss of a loved one, job loss), chronic illness (including psychological disorders within the past two years, or of schizophrenia, psychotic disorder, bipolar disorder, substance dependency, or personality disorder at any time in life), the use of hormonal contraceptives or other medication influencing the HPA axis were excluded (Kirschbaum et al., 1999). To reduce women-within variability and to assure comparability with male cortisol stress responses (Kirschbaum et al., 1999), testing days for female participants were scheduled according to their self-reported luteal phase (Kajantie and Phillips, 2006). On the testing day, female participants were asked to confirm their current menstrual cycle status, resulting in 95 women in their luteal (49 observers, 46 targets), 4 in their follicular (2 observers, 2 targets), and 9 in their menstrual phases (3 observers, 6 targets; see Table S1 for further sample characteristics).

##### Supplementary state stress measures

The Affect Grid (Russell et al., 1989) and a single-item, seven-point stress scale ("How stressed do you feel at this moment?"; ranging from "not at all" to "a lot") were also used to assess subjective stress experience in targets and observers. The Affect Grid is a visual single item scale that assesses the dimensions of pleasure-displeasure and arousal-sleepiness. These dimensions are aligned on the x- and y-axes of a grid, in which participants mark their current state. Due to the coverage of many stress-related feelings in the STAI and its frequent use, we consider the STAI state scale our main measure of subjective stress. Affect Grid and the single item stress scale were treated as exploratory variables and are reported on only in the Supplemental Material.

**Supplementary state and trait measures**

Observer's trait characteristics of empathy and compassion were assessed with the German version of the Interpersonal Reactivity Index (IRI; Davis, 1983), the Saarbrücker Persönlichkeitsfragebogen (Paulus, 2009). To capture trait power, the Generalized Sense of Power Scale (Anderson et al., 2012; Anderson and Galinsky, 2006) was employed. Further, observer's state empathy and compassion, using the Emotional Response Scale (ERS) (Batson et al., 1997), and observer's state power, using a self-created adjective list, were measured. In addition, chronic stress exposure of observers was operationalized with the Perceived Stress Scale (PSS) (Cohen et al., 1983). These questionnaires have been analysed and reported in detail in Heilmann et al. (2024) and are not part of the current study.

**Supplementary statistical analyses**

Initially, the current analyses were planned as a joint project with those published in (Heilmann et al., 2024), and preregistered together (https://osf.io/nbspw). However, to not overload the resulting manuscript, we decided to publish the oxytocin and testosterone data separately. While this naturally leads to some deviation, we stayed as close to the preregistration as possible. In detail, other than preregistered, we decided to include the manipulation (grouping) variable as a control variable and focus on the associations of empathic stress with oxytocin and testosterone independent of group belonging. This decision was made because of multicollinearity in our analyses. By including the grouping variable as a control variable, we were able to reduce multicollinearity (all variation inflation factors < 1.5). A further deviation was that we added a LMM examining the time course of oxytocin and testosterone to decide whether to use an AUCg or a change (reactivity) score for the main analyses.

Outliers were winsorized to 3 *SD*s from the mean. Random, single missings in the subjective and physiological stress data were replaced by the mean of previous and subsequent samples, if these were available. If one or two items of a multi-item questionnaire were missing, the mean of the respective questionnaire or subscale replaced the missing items (see Table S2 and S3 for the number of missing replacements). To facilitate interpretation and to provide standardized coefficients in the linear mixed models (LMM), continuous predictors were z-standardized instead of mean-centered (as initially preregistered).

Observer heart rate and HF-HRV baseline levels (at -31 min relative to stressor onset) were elevated compared to the subsequent measurement at -14 min and to target baseline levels (see Figures 3C and 3D in Heilmann et al., 2024), possibly due to a subtle stress response to the laboratory environment and the upcoming testing. As empathic stress effects are expected to be small overall, and staying congruent with our analysis approach in Heilmann et al. (2024), we excluded the observers' first sample at -31 min. The subsequent measure at -14 minutes was used as baseline in the LMMs instead.

*Model building for examining the time course of oxytocin and testosterone release*

The final model (Model 1), including a random intercept for the individual, was specified according to Raudenbush and Bryk's notation (Raudenbush and Bryk, 2002):

Model 1

Level 1: Oxytocin/testosterone$_{ti}$ = $\pi_{0i}$ + $\pi_{1i}$ (reactivity slope) + $\pi_{2i}$ (recovery slope) + $e_{ti}$

Level 2: $\pi_{0i}$ = $\beta_{00}$ + $\beta_{01}$ (sex female) + $\beta_{02}$ (empathy group) + $\beta_{03}$ (power group) + $\beta_{04}$ (further covariate) + $r_{0i}$

Fixed effects: Oxytocin/testosterone = $\gamma_{00}$ + $\gamma_{01}$ (sex female) + $\gamma_{02}$ (empathy group) + $\gamma_{03}$ (power group) + $\gamma_{04}$ (further covariate) + $\gamma_{10}$ (reactivity slope) + $\gamma_{20}$ (recovery slope)

Random effects: Oxytocin/testosterone$_i$ = $r_{0i}$ + $e_{ts}$

*Model building for examining the role of oxytocin and testosterone in empathic stress responses*

The final model (Model 2), including a random intercept for the individual, was specified according to Raudenbush and Bryk's notation (Raudenbush and Bryk, 2002):

Model 2

Level 1: Stress marker$_{ti}$ = $\pi_{0i} + \pi_{1i}$ (reactivity slope) + $e_{ti}$

Level 2: $\pi_{0i}$ = $\beta_{00}$ + $\beta_{01}$ (target stress reactivity) + $\beta_{02}$ (observer oxytocin/ testosterone reactivity) + $\beta_{03}$ (empathy group) + $\beta_{04}$ (power group) + $\beta_{05}$ (further covariate) + $r_{0i}$

Fixed effects: Stress marker = $\gamma_{00}$ + $\gamma_{01}$ (target stress reactivity) + $\gamma_{02}$ (observer oxytocin/ testosterone reactivity) + $\gamma_{03}$ (empathy group) + $\gamma_{04}$ (power group) + $\gamma_{05}$ (further covariate) + $\gamma_{10}$ (reactivity slope) + + $\gamma_{11}$ (reactivity slope* target stress reactivity) + $\gamma_{12}$ (reactivity slope*observer oxytocin/ testosterone reactivity) + $\gamma_{01}$ (target stress reactivity)*$\gamma_{02}$ (observer oxytocin/ testosterone reactivity) + $\gamma_{11}$ (reactivity slope* target stress reactivity)*$\gamma_{02}$ (observer oxytocin/ testosterone reactivity)

Random effects: Stress marker$_i$ = $r_{0i}$ + $e_{ts}$

*Exploratory subjective stress markers*

Exploratory analyses were conducted to examine associations of observers' oxytocin and testosterone reactivity with their subjective empathic stress responses in arousal, valence, and the stress scale (adding up to three models for each of testosterone and oxytocin). A continuous time variable was utilized to estimate the reactivity of each stress marker. The continuous time variable modelled the minutes between measurement time-points from baseline to peak (see Table S4 for the exact timeline of each stress marker). The intercept, defined as the measurement time point of observers' highest average peak, was set to 0. Contrasts were set to the predefined "contr.sum" in R, so that our intercept represented the grand mean of the sample. Next to the time slope, targets' stress reactivity (Δtarget), observers' oxytocin or testosterone reactivity (Δoxytocin or Δtestosterone), their interactions with the reactivity slope, and covariates (observer's sex and the grouping variable of the manipulation for all models) were included in the model (see previous paragraph for the final model building). ΔTarget, observer Δoxytocin, and Δtestosterone were operationalized as a change score from baseline to the individual peak. Considering the law of initial value for physiological markers (Wilder, 1957), observer Δoxytocin and Δtestosterone were adjusted for baseline levels by extracting the standardized change score residuals from a regression model.

#### Supplementary Results and Discussion

*The role of oxytocin and testosterone reactivity in empathic stress responses of the exploratory subjective stress markers*

No effects of observer oxytocin reactivity were found on vicarious (all $t \leq |1.20|$, all $p \geq .23$) or resonant stress (all $t \leq |1.60|$, all $p \geq .11$, Table S5) in any of the exploratory subjective stress markers.

Regarding testosterone, there were no associations between observer testosterone reactivity and vicarious (all $t \leq |0.83|$, all $p \geq .41$) or resonant stress responses in arousal, valence or the stress scale (all $t \leq |1.91|$, all $p \geq .06$, Table S6).

In sum, consistent with our findings in the STAI, empathic stress responses in the exploratory subjective stress markers were not related to observer oxytocin or testosterone reactivities. Thus, it seems that oxytocin and testosterone are not involved in the subjective empathic response. However, inaccuracy during the assessment might have influenced these results as self-reports are susceptible to bias, such as social desirability (Althubaiti, 2016).

**Table S1. Sample characteristics per group.**

| | **Targets ($n$ = 108)** | **Observer ($n$ = 108)** |
|---|---|---|
| | Number of participants | |
| Sex (male/female) | | |
| | 54/54 | 54/54 |
| Menstrual cycle status (follicular/luteal/menstrual phase) | | |
| | 2/46/6 | 2/49/3 |
| | Mean (*SD*) | |
| Age | 24.76 (4.79) | 24.74 (4.90) |
| BMI | 22.87 (2.67) | 23.03 (2.60) |

*Note.* BMI: Body mass index.

**Table S2. ECG time frames across experimental phases**

| Experimental phase | Time frame (relative to stressor onset) | Duration |
|---|---|---|
| Baseline | -35 min to 50 min | 6 min |
| Anticipation phase 1 | -14 min to -8 min | 6 min |
| Anticipation phase 2 | -8 min to -2 min | 6 min |
| Stress phase 1 | 0 min to 5 min | 5 min |
| Stress phase 2 | 5 min to 10 min | 5 min |
| Recovery phase 1 | 10 min to 15.5. min | 5.5 min |
| Recovery phase 2 | 15.5 min to 21. min | 5.5 min |
| Recovery phase 3 | 21 min to 26.5. min | 5.5 min |
| Recovery phase 4 | 26.5 min to 32 min | 5.5 min |
| Recovery phase 5 | 32 min to 37.5. min | 5.5 min |
| Recovery phase 6 | 37.5 min to 43 min | 5.5 min |
| Recovery phase 7 | 43 min to 48.5. min | 5.5 min |

*Note.* ECG time frames were extended by 30-60 sec, if possible, to ensure that each frame included at least 5 min of ECG recording after cutting artifacts (duration recommended for calculation of average time bins by the Task Force of the European Society and The North American Society of Pacing and Electrophysiology, 1996).

**Table S3. Number of participants with available data (and replacements/winsorized outliers) per measure and measurement time point.**

| *Targets and observers (n = 216)* | | | | |
|---|---|---|---|---|
| | **Cortisol** | **Heart Rate** | **HF-HRV** | **STAI** |
| Measurement time point | | | | |
| Sample1 | 215 (-/1) | 216 (-/3) | 216 (-/-) | 216 (-/1) |
| Sample2 | 216 (-/1) | 215 (-/3) | 215 (-/2) | 216 (2/-) |
| Sample3 | 216 (-/3) | 214 (-/2) | 214 (-/2) | 216 (-/-) |
| Sample4 | 216 (-/2) | 214 (-/1) | 214 (-/-) | 216 (-/-) |
| Sample5 | 216 (-/2) | 214 (-/2) | 214 (-/2) | 216 (1/1) |
| Sample6 | 216 (-/2) | 215 (-/1) | 215 (-/-) | 216 (-/-) |
| Sample7 | 216 (2/2) | 214 (-/1) | 214 (-/-) | 216 (-/2) |
| Sample8 | 216 (-/2) | 214 (-/2) | 214 (-/-) | 216 (-/2) |
| Sample9 | 216 (-/2) | 214 (-/1) | 214 (-/-) | 216 (-/3) |
| Sample10 | | 214 (-/1) | 214 (-/-) | |
| Sample11 | | 215 (-/2) | 215 (-/-) | |
| Sample12 | | 216 (-/-) | 216 (-/-) | |
| | **Stressscale** | **Valence** | **Arousal** | |
| Measurement time point | | | | |
| Sample1 | 214 (-/4) | 216 (-/-) | 216 (-/-) | |
| Sample2 | 214 (3/-) | 216 (1/-) | 216 (1/1) | |
| Sample3 | 215 (5/-) | 216 (1/-) | 216 (1/-) | |
| Sample4 | 215 (1/1) | 216 (-/-) | 216 (-/-) | |
| Sample5 | 215 (4/2) | 216 (2/-) | 216 (2/-) | |
| Sample6 | 215 (4/3) | 216 (-/1) | 216 (-/1) | |
| Sample7 | 214 (5/2) | 216 (-/2) | 216 (-/1) | |
| Sample8 | 215 (-/5) | 216 (-/4) | 216 (-/1) | |
| Sample9 | 215 (-/5) | 216 (-/4) | 216 (-/1) | |
| *Only in observers (n = 108)* | | | | |
| | **Oxytocin** | **Testosterone** | | |
| Measurement time point | | | | |
| Sample1 | 106 (-/-) | 108 (-/-) | | |
| Sample2 | 106 (-/-) | 108 (1/-) | | |
| Sample3 | 106 (-/-) | 108 (-/-) | | |
| Sample4 | 106 (2/-) | 108 (-/-) | | |
| Sample5 | 107 (-/-) | 108 (-/-) | | |

*Note.* Missing data are attributable to insufficient saliva volume (cortisol, oxytocin and testosterone), too much noise in the ECG data (heart rate and HF-HRV) and uncompleted questionnaires.
HF-HRV: High-frequency heart rate variability, STAI: State Trait Anxiety Inventory (Spielberger et al., 1983).

**Table S4. Number of possible and replaced data points within questionnaires per measurement time point.**

| | *N* possible data points | *N* replaced missing data points |
|---|---|---|
| **STAI (sample 1)** | 4320 (100%) | 12 (0.28%) |
| **STAI (sample 2)** | 4320 (100%) | 5 (0.12%) |
| **STAI (sample 3)** | 4320 (100%) | 4 (0.09%) |
| **STAI (sample 4)** | 4320 (100%) | 4 (0.09%) |
| **STAI (sample 5)** | 4320 (100%) | 4 (0.09%) |
| **STAI (sample 6)** | 4320 (100%) | 5 (0.12%) |
| **STAI (sample 7)** | 4320 (100%) | 8 (0.19%) |
| **STAI (sample 8)** | 4320 (100%) | 2 (0.05%) |
| **STAI (sample 9)** | 4320 (100%) | 5 (0.12%) |

*Note.* STAI: State Trait Anxiety Inventory (Spielberger et al., 1983).

**Table S5. Timeline of peak and reactivity slope in the models 2 for each stress marker.**

| Stress marker | Reactivity slope | Intercept (peak) |
|---|---|---|
| STAI | -25 min to 10 min | 10 min |
| Arousal | -25 min to 10 min | 10 min |
| Valence | -25 min to 10 min | 10 min |
| Stress scale | -25 min to 10 min | 10 min |
| Cortisol | -25 min to 15 min | 15 min |
| Heart rate | 2nd time frame (-14 to -8 min) to 5th time frame (5 to 10 min) | 5th time frame (5 to 10 min) |
| HF-HRV | 2nd time frame (-14 to -8 min) to 5th time frame (5 to 10 min) | 5th time frame (5 to 10 min) |

*Note.* Minutes are given relative to stressor onset. For heart rate and HF-HRV a mean was calculated over a specific time frame instead of one measurement time point as for cortisol and subjective stress markers.
STAI: State Trait Anxiety Inventory (Spielberger et al., 1983), HF-HRV: High-frequency heart rate variability.

**Table S6. A LMM examining associations between observers' oxytocin reactivity and empathic stress responses in the arousal (A) and valence (B) scale of the Affect Grid (Russell et al., 1989), and in a single-item stress scale (C).**

Predictor variables written in italics represent resonant stress responses. ΔTarget: target stress reactivity; Δoxytocin: observer oxytocin reactivity.

| **Fixed effects** | **A Arousal** | | | **B Valence** | | |
|---|---|---|---|---|---|---|
| | *β* | 95% CI | *t* (df) | *β* | 95% CI | *t* (df) |
| Intercept (peak) | 5.465 | 5.120; 5.810 | 30.36 (183)*** | 5.557 | 5.251; 5.864 | 34.79 (193)*** |
| Reactivity slope | 0.055 | 0.043; 0.067 | 8.97 (208)*** | -0.047 | -0.058; -0.036 | -8.32 (208)*** |
| *ΔTarget* | 0.285 | -0.061; 0.631 | 1.58 (182) | -0.063 | -0.381; 0.255 | -0.38 (186) |
| *Reactivity slope*Δtarget* | 0.005 | -0.007; 0.017 | 0.84 (208) | -0.002 | -0.013; 0.009 | -0.32 (208) |
| ΔOxytocin | -0.048 | -0.402; 0.305 | -0.26 (182) | 0.047 | -0.265; 0.359 | 0.29 (191) |
| Reactivity slope* Δoxytocin | -0.004 | -0.017; 0.008 | -0.71 (208) | 0.001 | -0.010; 0.012 | 0.16 (208) |
| *ΔTarget*Δoxytocin* | -0.240 | -0.572; 0.093 | -1.38 (179) | 0.225 | -0.074; 0.524 | 1.44 (192) |
| *Reactivity slope* Δtarget*Δoxytocin* | -0.006 | -0.017; 0.005 | -0.98 (208) | 0.009 | -0.002; 0.019 | 1.60 (208) |
| Sex | -0.078 | -0.374; 0.218 | -0.50 (99) | 0.172 | -0.090; 0.434 | 1.25 (99) |
| Group1 | 0.265 | -0.147; 0.678 | 1.23 (99) | -0.022 | -0.382; 0.338 | -0.12 (99) |
| Group2 | -0.377 | -0.788; 0.033 | -1.76 (99)† | 0.172 | -0.186; 0.531 | 0.92 (99) |
| **Random Effects** | | | | | | |
| | Variance *(SD)* | | | Variance *(SD)* | | |
| Individual | 1.60 (1.26) | | | 1.14 (1.07) | | |

| **Fixed effects** | **C Stress scale** | | |
|---|---|---|---|
| | *β* | 95% CI | *t* (df) |
| Intercept (peak) | 2.875 | 2.668; 3.082 | 26.63 (149)*** |
| Reactivity slope | 0.024 | 0.018; 0.030 | 7.79 (205)*** |
| *ΔTarget* | -0.085 | -0.305; 0.135 | -0.74 (146) |
| *Reactivity slope*Δtarget* | 0.001 | -0.005; 0.007 | 0.32 (204) |
| ΔOxytocin | 0.016 | -0.195; 0.226 | 0.14 (148) |
| Reactivity slope* Δoxytocin | -0.004 | -0.010; 0.002 | -1.20 (205) |
| *ΔTarget*Δoxytocin* | -0.009 | -0.162; 0.144 | -0.11 (147) |
| *Reactivity slope* Δtarget*Δoxytocin* | -0.002 | -0.006; 0.003 | -0.75 (204) |
| Sex | -0.027 | -0.213; 0.158 | -0.28 (98) |
| Group1 | 0.104 | -0.163; 0.371 | 0.74 (98) |
| Group2 | -0.336 | -0.601; -0.071 | -2.42 (98)* |
| **Random Effects** | | | |
| | Variance *(SD)* | | |
| Individual | 0.77 (0.88) | | |

*Note.* Satterthwaite approximation for degrees of freedom. Models based on 318 observations from 106 dyads (A, arousal), 318 observations from 106 dyads (B, valence), and 313 observations from 105 dyads (C, stress scale). Reactivity slope = variable modelling the time from baseline to peak; *β* = standardized beta coefficients; CI = confidence interval. Contrasts were set to the predefined "contr.sum" in R, so that the intercept (peak) represented the grand mean.

Data was winsorized to 3 *SDs* to the mean. Oxytocin data was additionally logerithmized. Age, target stress and observer reactivity were z-standardized.
† $p < .10$, * $p < .05$, ** $p < .01$, *** $p < .001$.

**Table S7. A LMM examining associations between observers' testosterone reactivity and empathic stress responses in the arousal (A) and valence (B) scale of the Affect Grid (Russell et al., 1989), and in a single-item stress scale (C).**

Predictor variables written in italics represent resonant stress responses. ΔTarget: target stress reactivity; Δtestosterone: observer testosterone reactivity.

| Fixed effects | A Arousal | | | B Valence | | |
|---|---|---|---|---|---|---|
| | *β* | 95% CI | *t* (df) | *β* | 95% CI | *t* (df) |
| Intercept (peak) | 5.461 | 5.116; 5.807 | 30.33 (187)*** | 5.600 | 5.293; 5.906 | 35.06 (199)*** |
| Reactivity slope | 0.053 | 0.041; 0.065 | 8.74 (212)*** | -0.046 | -0.057; -0.034 | -8.01 (212)*** |
| *ΔTarget* | 0.265 | -0.105; 0.587 | 1.45 (187) | -0.047 | -0.366; 0.272 | -0.28 (193) |
| *Reactivity slope*Δtarget* | 0.005 | -0.007; 0.017 | 0.81 (212) | -0.001 | -0.012; 0.010 | -0.21 (212) |
| ΔTestosterone | -0.035 | -0.409; 0.340 | -0.18 (183) | 0.086 | -0.231; 0.403 | 0.52 (194) |
| Reactivity slope* Δtestosterone | -0.005 | -0.018; 0.007 | -0.83 (212) | 0.005 | -0.006; 0.016 | 0.82 (212) |
| *ΔTarget*Δtestosterone* | 0.006 | -0.340; 0.412 | 0.03 (183) | 0.257 | -0.001; 0.515 | 1.91 (198)† |
| *Reactivity slope* Δtarget*Δtestosterone* | 0.003 | -0.011; 0.016 | 0.36 (212) | 0.007 | -0.002; 0.017 | 1.54 (212) |
| Sex | -0.032 | -0.330; 0.265 | -0.21 (101) | 0.168 | -0.095; 0.430 | 1.22 (101) |
| Group1 | 0.222 | -0.195; 0.639 | 1.02 (101) | -0.003 | -0.354; 0.348 | -0.02 (101) |
| Group2 | -0.363 | -0.776; 0.050 | -1.68 (101) | 0.169 | -0.186; 0.523 | 0.91 (101) |
| **Random Effects** | | | | | | |
| | Variance *(SD)* | | | Variance *(SD)* | | |
| Individual | 1.60 (1.27) | | | 1.10 (1.05) | | |

| Fixed effects | C Stress scale | | |
|---|---|---|---|
| | *β* | 95% CI | *t* (df) |
| Intercept (peak) | 2.864 | 2.658; 3.069 | 26.66 (153)*** |
| Reactivity slope | 0.023 | 0.017; 0.029 | 7.67 (209)*** |
| *ΔTarget* | -0.119 | -0.342; 0.104 | -1.02 (150) |
| *Reactivity slope*Δtarget* | 0.002 | -0.004; 0.008 | 0.63 (208) |
| ΔTestosterone | -0.036 | -0.251; 0.179 | -0.32 (148) |
| Reactivity slope* Δtestosterone | -0.002 | -0.008; 0.004 | -0.73 (208) |
| *ΔTarget*Δtestosterone* | 0.070 | -0.077; 0.218 | 0.92 (152) |
| *Reactivity slope* Δtarget*Δtestosterone* | -0.001 | -0.006 0.003 | -0.62 (208) |
| Sex | -0.010 | -0.201; 0.181 | -0.10 (100) |
| Group1 | 0.064 | -0.200; 0.328 | 0.46 (100) |
| Group2 | -0.347 | -0.612; -0.082 | -2.50 (100)* |
| **Random Effects** | | | |
| | Variance *(SD)* | | |
| Individual | 0.77 (0.88) | | |

*Note.* Satterthwaite approximation for degrees of freedom. Models based on 324 observations from 108 dyads (A, arousal), 324 observations from 108 dyads (B, valence), and 319 observations from 107 dyads (C, stress scale). Reactivity slope = variable modelling the time from baseline to peak; *β* = standardized beta coefficients; CI = Confidence interval. Contrasts were set to the predefined "contr.sum" in R, so that the intercept (peak) represented the grand mean.

Data was winsorized to 3 *SDs* to the mean. Testosterone data was additionally logerithmized. Age, target stress and observer reactivity were z-standardized.
† $p < .10$, * $p < .05$, ** $p < .01$, *** $p < .001$.
